# Low Antibody Titers Demonstrate Significantly Increased Rate of SARS-CoV-2 Infection in a Highly Vaccinated Population from the National Basketball Association

**DOI:** 10.1101/2023.02.10.23285780

**Authors:** Caroline G. Tai, Miriam J. Haviland, Steven M. Kissler, Rachel M. Lucia, Michael Merson, Lisa L. Maragakis, David D. Ho, Deverick J. Anderson, John DiFiori, Nathan D. Grubaugh, Yonatan H. Grad, Christina DeFilippo Mack

## Abstract

SARS-CoV-2 antibody titers may serve as a correlate for immunity and could inform optimal booster timing. The relationship between antibody titer and protection from infection was evaluated in 2,323 vaccinated individuals from the National Basketball Association who had antibody levels measured from 9/12/2021—12/31/2021. Cox-proportional hazards models were used to estimate risk of infection within 90-days of serologic testing by titer level (<250, 250-800, and >800 AU/mL) and individuals were censored on date of booster receipt. The cohort was 78.2% male, 68.1% aged ≤ 40 years, and 56.4% vaccinated (primary series) with the Pfizer-BioNTech mRNA vaccine. Among the 2,248 individuals not yet boosted at testing, those with titers <250 AU/mL (adj HR: 2.4; 95% CI: 1.5, 3.7) and 250-800 800 AU/mL (adj HR: 1.5; 95% CI: 0.98, 2.4) had greater infection risk compared to those with titers >800 AU/mL. Serologic titers could inform individual COVID-19 risk and booster scheduling.

## Introduction

The effectiveness of COVID-19 vaccination for preventing SARS-CoV-2 infection wanes [1,2], and although additional vaccine doses reduce the risk of incident SARS-CoV-2 infection [3], the optimal schedule for these booster doses remains unclear [4–6]. Additionally, many factors—such as age, comorbidity and history of SARS-CoV-2 infection—impact immune response to SARS-CoV-2, suggesting that, although a one-size-fits-all time-based booster schedule may be the only practical approach currently, such a schedule ignores important individual-level heterogeneity [7,8]. Identifying an objective correlate of immunity, could help health care providers understand infection risk and the optimal booster timing to protect against SARS-CoV-2 infection.

SARS-CoV-2 antibody titers are a potential correlate of immunity that can be measured through serologic tests. Several of these tests have received Emergency Use Authorization from the Food and Drug Administration (FDA) since March 2020 [9], but none yet have full FDA approval to quantify immunity to SARS-CoV-2 infection, and it is possible that new tests are currently needed to account for the continuing evolution of SARS-CoV-2 variants. The FDA has indicated that more research is needed, and antibody testing is not currently recommended to assess a person’s level of immunity after COVID-19 vaccination [10]. While high SARS-CoV-2 antibody titers up to four weeks after primary series vaccinations are associated with decreased infection risk [11,12], there remains a gap in the understanding of antibody titer trajectories over longer periods of time since vaccination.

This study aimed to measure the relationship between antibody titers and incident SARS-CoV-2 infection among a cohort of individuals who received all doses of a SARS-CoV-2 primary vaccine series (hereafter referred to as vaccinated) and to evaluate the utility of serologic testing to estimate SARS-CoV-2 risk prior to receiving a booster. This was accomplished by 1) assessing the association between antibody titers and factors including time since vaccination, vaccine type, booster status, and prior SARS-CoV-2 infection, and 2) evaluating the relationship between antibody titers and rate of subsequent SARS-CoV-2 infection.

## Methods

Before the start of the 2021-2022 National Basketball Association (NBA) season, players and team staff underwent assessment of their SARS-CoV-2 antibody titers using the DiaSorin LIAISON SARS-CoV-2 TrimericS IgG assay (the “TrimericS Assay”) which was developed to measure antibodies against the WA-1 spike protein and received FDA emergency use authorization on May 19, 2021 [13]. At the time, the TrimericS assay was chosen for its high correlation to neutralizing antibodies compared to other commercially available platforms [14–17]. Individuals were included if they met the following criteria: aged 18 and older who voluntarily had their SARS-CoV-2 antibody titers measured using the TrimericS Assay from September 12, 2021 through December 31, 2021 and completed their primary SARS-CoV-2 vaccine series by time of antibody testing. Those who received a vaccine dose (primary or booster) or had a confirmed COVID-19 infection in the 14 days before their antibody test were excluded. If an individual had more than one antibody test during the study period, only the first test was included in these analyses. Relevant demographic and clinical characteristics (age, time since receipt of primary vaccine, and prior SARS-CoV-2 infection) were collected as part of the NBA occupational health program, as described [3,18,19].

The Advarra institutional review board determined the study met criteria for exemption status. Individuals signed health information authorizations allowing collection, storage, and use of health information by the NBA for monitoring purposes, including disclosure to medical experts.

### Antibody Titers Assessment

The TrimericS assay reports qualitative results (detected and not detected) and quantitative values in Arbitrary Units (AU) per milliliter (mL) when antibody titers are detected. Titer values ≥13 AU/mL are considered detectable, however only values between 13-800 AU/mL are reported by the assay while titers above this range are reported as >800 AU/mL [20] and could represent values substantially greater than 800 AU/mL. Individuals were categorized into three groups based on titer level: <250, 250-800, and >800 AU/mL. These cut-offs were chosen based on correlation between TrimericS assay values and virus neutralization assays for wildtype and Delta SARS-CoV-2 strains [21–24]. These assays showed that neutralizing titers of 100 were correlated with a 50% protective neutralization level for wildtype [23]. This corresponded to a cutoff of ≥189.09 TrimericS AU/mL (95% CI: 147.61-235.75) as described previously [24]. The cutoff of 250 was taken as a conservative upper bound to denote a group with potentially higher risk of SARS-CoV-2 infection.

### SARS-CoV-2 Infection

The primary outcome of interest was defined as a SARS-CoV-2 infection within 90 days following antibody measurement. Any individual with at least one positive test and clinical confirmation was defined as having a confirmed incident SARS-CoV-2 infection, as described [18].

When possible, at least one positive PCR sample from each confirmed case was sent for genomic sequencing. In some cases, SARS-CoV-2 sequencing was unsuccessful due to inadequate sample volume or low viral load. In these scenarios, cases before 12/3/2021 were assumed to be Delta and those on or after 2/1/2022 to be Omicron. Cases occurring between 12/3/2021 through 1/31/2022 when both variants were circulating in this cohort were considered not sequenced.

### Statistical Analysis

Antibody titers were summarized by demographic and clinical factors including vaccine type, booster status, and days since last dose of primary series. To evaluate the association between antibody titers and risk of subsequent incident SARS-CoV-2 infection, a time-to-event analysis was conducted among the subset who had not yet received a booster at the time of serologic testing. Rates of time to SARS-CoV-2 infection were estimated using life tables, Kaplan-Meier curves, and Cox proportional hazards regression models (with 95% confidence intervals). To assess the proportional hazards assumptions, plots of the survival function and log(-log(survival)) versus log(time) and models with time-dependent covariates for each independent variable of interest were evaluated. The risk of SARS-CoV-2 infection within 90 days of antibody measurement among the <250 AU/mL and 250-800 AU/mL titer groups were compared to the >800 AU/mL group (reference), overall and stratified by primary vaccine type. Individuals who received a booster within 90 days after serologic testing were censored on the date of their booster. Models were adjusted for age (years), months since last vaccine dose, and SARS-CoV-2 infection before antibody measurement (yes and no/unknown).

The predominant SARS-CoV-2 variant transitioned from Delta (B.1.617.2 and AY lineages) to Omicron (initially B.1.1.529, BA.1, and BA.1.1) in late November/December of 2021, during the follow-up period; therefore, sensitivity analyses were conducted to estimate variant-specific hazard ratios for Delta and Omicron infections, modeling Omicron (or Delta) as competing risks. Infections that could not be sequenced were also modeled as a competing risk. To evaluate how the estimated association between antibody titers and risk of SARS-CoV-2 infection may have been influenced by the selection of 90 days for the follow-up period, a sensitivity analysis using a 60-day follow-up period was also conducted. As in the primary analyses, individuals who received a booster during the follow-up period were censored on date of receipt. All analyses were conducted using SAS Enterprise Guide version 8.2 and R version 4.2.0.

## Results

From September 12, 2021 through December 31, 2021, a total of 2,388 vaccinated adults underwent antibody testing via the TrimericS Assay. Of these, 54 (2.3%) received a primary vaccine or booster dose, and 11 (0.5%) had a confirmed SARS-CoV-2 infection in the 14 days before their serologic test and were excluded. This resulted in a cohort of 2,323 individuals. Antibody titers for 92% of the cohort were measured during September 12, 2021 to October 12, 2021. The cohort was 78.2% male, 68.1% were aged ≤ 40 years, and 56.4% were vaccinated (primary series) with the Pfizer-BioNTech mRNA vaccine (Table 1). While 75 (3.2%) individuals had received a SARS-CoV-2 booster (monovalent) by the time of serologic testing, 1,934 (86.0%) individuals who were unboosted at serologic testing later received a booster within the 90 day follow up period. Most individuals in this analysis were several months out from their primary vaccination series: 1,735 (77.2%) of the vaccinated unboosted individuals were 4-6 months post-vaccination at antibody measurement, and 58 (77.3%) of the boosted individuals were more than 7 months primary post-vaccination.

**Table 1:** Characteristics at time of serologic test.

| Characteristics | Overall<br>n=2,323 | Vaccinated* unboosted at<br>serologic test<br>n=2,248 | Boosted at<br>serologic test<br>n=75 |
| --- | --- | --- | --- |
| **Sex, n (%)** |  |  |  |
| Female | 218 (9.4) | 211 (9.4) | 7 (9.3) |
| Male | 1,817 (78.2) | 1,754 (78.0) | 63 (84.0) |
| Not reported | 288 (12.4) | 283 (12.6) | 5 (6.7) |
| **Age in years, n (%)** |  |  |  |
| 18-30 | 926 (39.9) | 915 (40.7) | 11 (14.7) |
| 31-40 | 654 (28.2) | 637 (28.3) | 17 (22.7) |
| 41-50 | 409 (17.6) | 389 (17.3) | 20 (26.7) |
| 51-60 | 245 (10.5) | 230 (10.2) | 15 (20.0) |
| ≥ 61 | 89 (3.8) | 77 (3.4) | 12 (16.0) |
| **Time since last dose of primary vaccine series, n (%)** |  |  |  |
| < 4 months | 285 (12.3) | 284 (12.6) | 1 (1.3) |
| 4-6 months | 1,751 (75.4) | 1,735 (77.2) | 16 (21.3) |
| ≥ 7 months | 287 (12.4) | 229 (10.2) | 58 (77.3) |
| **SARS-CoV-2 infection before serologic test, n (%)**<sup>†</sup> |  |  |  |
| Infection at any timepoint before serologic test | 390 (16.8) | 388 (17.3) | 2 (2.7) |
| Infection before last dose of primary series | 290 (74.4) | 289 (74.5) | 1 (50.0) |
| Infection after last dose of primary series | 100 (25.6) | 99 (25.5) | 1 (50.0) |
| **Primary vaccine series type, n (%)** |  |  |  |
| Vector - J&J/Janssen | 493 (21.2) | 483 (21.5) | 10 (13.3) |
| mRNA - Moderna | 520 (22.4) | 504 (22.4) | 16 (21.3) |
| mRNA - Pfizer-BioNTech | 1,310 (56.4) | 1,261 (56.1) | 49 (65.3) |
| **Qualitative antibody test result, n (%)** |  |  |  |
| Detected | 2,262 (97.4) | 2,187 (97.3) | 75 (100.0) |
| Not detected | 61 (2.6) | 61 (2.7) | 0 (0.0) |

Low Antibody Titer Increases SARS-COV-2 Rate
| Antibody titer category, n (%) |  |  |  |
| --- | --- | --- | --- |
| >800 AU/mL | 594 (25.6) | 524 (23.3) | 70 (93.3) |
| 250-800 AU/mL | 714 (30.7) | 709 (31.5) | 5 (6.7) |
| <250 AU/mL ‡ | 1,015 (43.7) | 1,015 (45.2) | 0 (0.0) |
| Antibody titer, AU/mL |  |  |  |
| Median | 309.0 | 293.5 | >800.0 |
| IQR: 25<sup>th</sup>, 75<sup>th</sup> percentile | 125.0, >800.0 | 121.0, 740.5 | Not Available<sup>§</sup> |

\* An individual is considered Vaccinated if they have received all doses of a SARS-CoV-2 primary vaccine series and at least 14 days have passed since they received the last dose. For example, if the final primary series dose was June 1, 2021 then the individual would be considered vaccinated on June 16, 2021.
† Individuals with at least one positive test and clinical confirmation from March 2020 through 14 days before their serologic test were considered to have a prior SARS-CoV-2 infection. Individuals with a SARS-CoV-2 infection within the 14 days before their serologic test were excluded from the analytic cohort.
‡ Includes “not detected” results, which were reported as a titer value of “<13 AU/mL”.
§ When titers were described as a continuous variable, values >800 were treated as 800 AU/mL and those with values <13 as 13 AU/mL. The IQR is “Not Available” when the 25th and 75th percentile were >800.0 AU/mL since these values exist beyond the reportable limit of detection authorized by the FDA for the TrimericS assay

There were 2,248 vaccinated individuals with detected titer values who were not yet boosted. Among these, the median antibody titer was 293.5 AU/mL, with a wide interquartile range (IQR): 121.0, 740.5. Titers were highest among those within 4 months of their primary series (Median: >800 AU/mL; IQR: 335.5, >800) and lower among individuals who had completed their primary series >7 months before their serologic test (Median: 141.0 AU/mL; IQR: 71.4, 338.0; Figure 1A). Titers varied by vaccine type, with J&J/Janssen vaccine recipients having lower titers (Median: 95.8 AU/mL; IQR: 24.5, 438.0) than Moderna and Pfizer-BioNTech vaccine recipients (Median: 463.0 AU/mL; IQR: 256.5, >800.0 and Median: 285.0 AU/mL; IQR: 138.0, 748.0, respectively; Table 2). Even among the individuals vaccinated <4 months before antibody measurement, J&J recipients had a lower median titer of 114 AU/mL (IQR: 26.2, 800.0) compared to mRNA recipients who had a median titer of >800 AU/mL (Figures 1B-1D, Table 2). This was also reflected in the qualitative results where 54 (88.5%) out of the 61 people with negative (“Not Detected”) results were J&J/Janssen primary vaccine recipients (Table 1). The 75 people who were boosted by the time of their serologic test had the highest titers, at a median of >800.0 AU/mL, with no variation across clinical characteristics of interest (Table 2).

**Figure 1:**
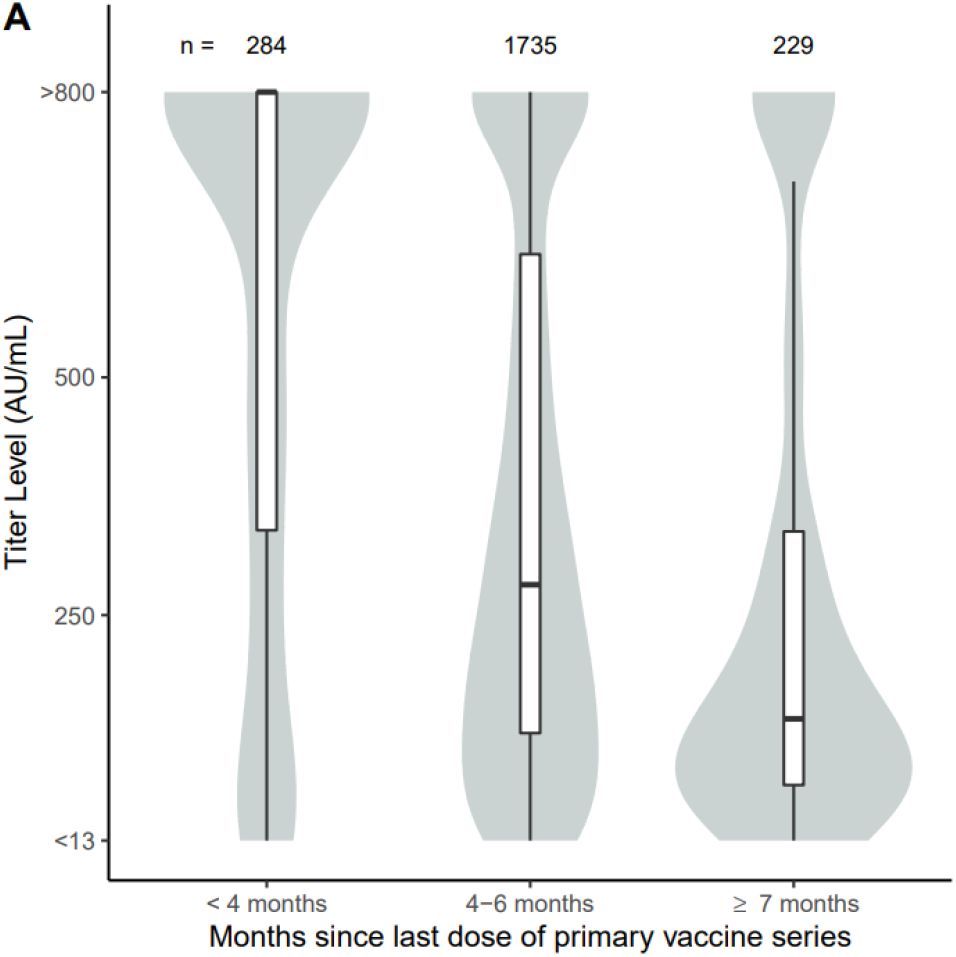
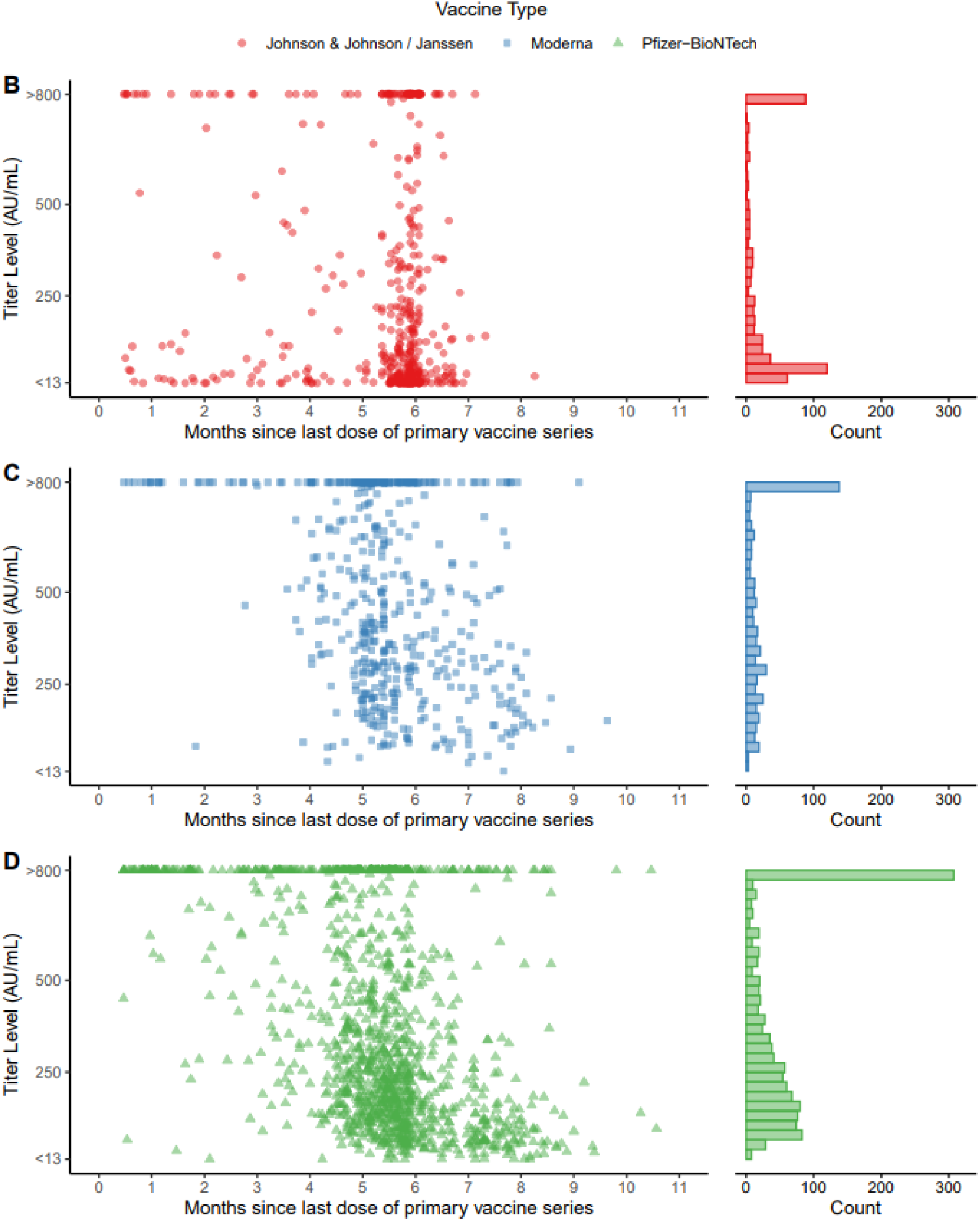
Antibody titers by primary series vaccine type and time since vaccination. Data in Figures 1A - D are restricted to individuals who were vaccinated unboosted at time of serologic test. Titer values >800 were treated as 800 AU/mL and those <13 as 13 AU/mL. Figure 1A shows the distribution of titer values by months since completion of the primary vaccine series. Values in the box and whisker plots should be interpreted as follows: bottom and top of box are respectively the 25^th^ and 75^th^ percentile, the center bar is the median (50^th^ percentile), and whiskers are 1.5 times the interquartile range (25^th^ - 75^th^ percentile). The shaded area shows the kernel density estimate at each titer value. Figures 1B-D show the distribution of titer values by month since completion of the primary vaccine series, stratified by primary series type. A histogram of the number of individuals is presented alongside each scatterplot to illustrate density of individuals by titer value.

**Table 2:** Antibody titer value* (AU/mL) by vaccine primary series type.

| Characteristic | Vaccinated unboosted at serologic test |  |  |  |  |  |  |  |  | Boosted at serologic Test |  |
| --- | --- | --- | --- | --- | --- | --- | --- | --- | --- | --- | --- |
|  | J&J/Janssen |  |  | Moderna |  |  | Pfizer-BioNTech |  |  |  |  |
|  | n (%) | Median | IQR | n (%) | Median | IQR | n (%) | Median | IQR | n (%) | Median <sup>‡</sup> |
| Overall | 483 | 95.8 | 24.5, 438.0 | 504 | 463.0 | 256.5, >800.0 | 1,261 | 285.0 | 138.0, 748.0 | 75 | >800.0 |
| Age |  |  |  |  |  |  |  |  |  |  |  |
| 18-40 | 376 (77.8) | 117.0 | 31.5, 462.0 | 304 (60.3) | 558.0 | 336.5, >800.0 | 872 (69.2) | 356.5 | 180.5, >800.0 | 28 (37.3) | >800.0 |
| 41-60 | 100 (20.7) | 32.8 | 13.4, 342.5 | 177 (35.1) | 309.0 | 183.0, 576.0 | 342 (27.1) | 174.0 | 87.2, 425.0 | 35 (46.7) | >800.0 |
| ≥ 61 | 7 (1.4) | 28.5 | <13.0, 196.0 | 23 (4.6) | 243.0 | 103.0, 638.0 | 47 (3.7) | 90.1 | 33.5, 214.0 | 12 (16.0) | >800.0 |
| Time since last primary series dose |  |  |  |  |  |  |  |  |  |  |  |
| < 4 months | 73 (15.1) | 114.0 | 26.2, >800.0 | 38 (7.5) | >800.0 | 791.0, >800.0 | 173 (13.7) | >800.0 | 527.0, >800.0 | 1 (1.3) | >800.0 |
| 4-6 months | 406 (84.1) | 91.7 | 23.2, 365.0 | 396 (78.6) | 463.0 | 282.0, 800.0 | 933 (74.0) | 267.0 | 145.0, 578.0 | 16 (21.3) | >800.0 |
| ≥ 7 months | 4 (0.8) | 138.0 | 83.5, 471.0 | 70 (13.9) | 241.5 | 139.0, 492.0 | 155 (12.3) | 105.0 | 59.7, 260.0 | 58 (77.3) | >800.0 |
| Time between serologic test and previous SARS-CoV-2 infection <sup>†</sup> |  |  |  |  |  |  |  |  |  |  |  |
| No known prior infection | 379 (78.5) | 49.0 | 19.0, 285.0 | 438 (86.9) | 413.5 | 242.0, 768.0 | 1,043 (82.7) | 234.0 | 121.0, 538.0 | 73 (97.3) | >800.0 |
| 0-6 months | 33 (6.8) | >800.0 | 349.0, >800.0 | 20 (4.0) | >800.0 | 508.0, >800.0 | 62 (4.9) | >800.0 | Not Available* | 1 (1.3) | >800.0 |
| ≥ 6 months | 71 (14.7) | 321.0 | 189.0, 627.0 | 46 (9.1) | >800.0 | 504.0, >800.0 | 156 (12.4) | 714.0 | 363.5, >800.0 | 1 (1.3) | >800.0 |

IQR: interquartile range
\*When titers were described as a continuous variable, values >800 were treated as 800 AU/mL and those with values <13 as 13 AU/mL. The IQR is “Not Available” when the 25th and 75th percentile were >800.0 AU/mL since these values exist beyond the reportable limit of detection authorized by the FDA for the TrimericS assay.
†Individuals with at least one positive test and clinical confirmation from March 2020 through 14 days before their serologic test were considered to have a prior SARS-CoV-2 infection. Individuals with a SARS-CoV-2 infection within the 14 days before their serologic test were excluded from the analytic cohort.
‡The interquartile range was not reported for boosted individuals as all values for the 25<sup>th</sup> and 75<sup>th</sup> percentiles were beyond the reportable limit of detection (>800 AU/mL) authorized by the FDA for the TrimericS assay. The range of titer values among individuals boosted at the time of serologic testing was 628.0 to >800.0 AU/mL; 93% of individuals boosted before serologic testing had titers >800.0 AU.

In total, 136 (6.0%) individuals had a confirmed SARS-CoV-2 infection within 90 days following serologic testing and before receiving a booster, 1,934 (86.0%) did not experience an infection before being censored at the time of booster receipt, and 178 (7.9%) did not have an infection or receive a booster during the follow-up period. Among those censored due to booster receipt, the median follow-up time was 49 days (IQR: 38, 61) with 338 (17.5%) censored within the first 30 days. The proportion of individuals who experienced a SARS-CoV-2 infection over the 90-day follow-up period was lower among those with titers <250 AU/mL than those with titers >800 AU/mL (5.5% vs 8.6%). Infections among the 1,015 individuals with low titers, however, tended to occur earlier in follow-up: 67.3% of infections among individuals with titers <250 AU/mL (compared to 26.7% of individuals with titers >800 AU/mL) occurred prior to day 70 (Figure 2, Table 3), thus the rate of infection among this group was 2.4 (95% CI: 1.5, 3.7) times higher than among those with >800 AU/mL, after adjusting for age, time since last primary vaccine dose, and SARS-CoV-2 infection. The day 70 timepoint coincided with the start of the Omicron wave in the United States (US) for most individuals. The 709 individuals with titers of 250-800 AU/mL had an elevated, although not significantly different relative rate of infection compared to the >800 AU/mL group (adj HR: 1.5; 95% CI: 0.98, 2.4). Similar patterns were observed when stratified by primary vaccine type (Table 4). Among the 1,261 Pfizer-BioNTech recipients, those with titers <250 AU/mL had lower cumulative incidence of SARS-CoV-2 infection but were more likely to have infections early in follow-up and therefore a greater rate of infection compared to those >800 AU/mL (adj HR: 2.0; 95% CI: 1.1, 3.6; Table 4). Those with titers of 250-800 AU/mL also had a lower cumulative incidence of infection, but a faster rate of infection relative to those with >800 AU/mL (adj HR: 1.8; 95% CI: 1.1, 2.9). Though not statistically significant, similar findings were observed among Moderna recipients (Table 4).

**Table 3:** Life table for cumulative incidence of SARS-CoV-2 infection by antibody titer category.

| Time<br>Interval<br>(days) | Antibody titer category |  |  |  |  |  |  |  |  |  |  |  |  |  |  |
| --- | --- | --- | --- | --- | --- | --- | --- | --- | --- | --- | --- | --- | --- | --- | --- |
|  | <250 AU/mL |  |  |  |  | 250-800 AU/mL |  |  |  |  | >800 AU/mL |  |  |  |  |
|  | Number of<br>individuals | Number of<br>individuals | Effective<br>sample<br>size* | Interval<br>incidence<br>rate | Cumulative<br>incidence<br>rate | Number of<br>individuals | Number of<br>individuals | Effective<br>sample<br>size* | Interval<br>incidence<br>rate | Cumulative<br>incidence<br>rate | Number of<br>individuals | Number of<br>individuals | Effective<br>sample<br>size* | Interval<br>incidence<br>rate | Cumulative<br>incidence<br>rate |
|  | infected<br>n=52 | censored<br>n=963 |  |  |  | infected<br>n=39 | censored<br>n=970 |  |  |  | infected<br>n=45 | censored<br>n=479 |  |  |  |
| 0 | 0 | 0 | 1015 | 0.000 | 0.000 | 0 | 0 | 709 | 0.000 | 0.000 | 0 | 0 | 524 | 0.000 | 0.000 |
| 1, 10 | 1 | 64 | 983 | 0.001 | 0.000 | 0 | 15 | 701.5 | 0.000 | 0.000 | 0 | 7 | 520.5 | 0.000 | 0.000 |
| 11, 20 | 3 | 79 | 910.5 | 0.003 | 0.001 | 1 | 39 | 674.5 | 0.001 | 0.000 | 0 | 21 | 506.5 | 0.000 | 0.000 |
| 21, 30 | 2 | 78 | 829 | 0.002 | 0.004 | 2 | 22 | 643 | 0.003 | 0.001 | 2 | 13 | 489.5 | 0.004 | 0.000 |
| 31, 40 | 15 | 128 | 724 | 0.021 | 0.007 | 2 | 70 | 595 | 0.003 | 0.005 | 2 | 20 | 471 | 0.004 | 0.004 |
| 41, 50 | 8 | 258 | 516 | 0.016 | 0.027 | 3 | 185 | 465.5 | 0.006 | 0.008 | 0 | 85 | 416.5 | 0.000 | 0.008 |
| 51, 60 | 1 | 161 | 298.5 | 0.003 | 0.042 | 1 | 128 | 306 | 0.003 | 0.014 | 4 | 77 | 335.5 | 0.012 | 0.008 |
| 61, 70 | 5 | 90 | 172 | 0.029 | 0.046 | 3 | 88 | 197 | 0.015 | 0.018 | 4 | 47 | 269.5 | 0.015 | 0.020 |
| 71, 80 | 4 | 61 | 91.5 | 0.044 | 0.073 | 8 | 65 | 117.5 | 0.068 | 0.033 | 12 | 73 | 205.5 | 0.058 | 0.035 |
| 81, 90 | 13 | 44 | 35 | 0.371 | 0.114 | 19 | 58 | 48 | 0.396 | 0.098 | 21 | 136 | 89 | 0.236 | 0.091 |

\*Censored individuals are assumed to drop out of the risk set halfway through the interval, thus the effective sample size equals the number of individuals in the risk set at the start of the interval minus half of the number of individuals censored during the interval.

**Table 4:** Rate for SARS-CoV-2 infection by antibody titer, stratified by primary vaccine type.

|  | Antibody titer category |  |  |
| --- | --- | --- | --- |
|  | <250 AU/mL | 250-800 AU/mL | >800 AU/mL |
| **Overall** |  |  |  |
| n | 1,015 | 709 | 524 |
| Number (%) infected | 52 (5.1) | 39 (5.5) | 45 (8.6) |
| Crude HR (95% CI) | 2.1 (1.4, 3.1) | 1.4 (0.93, 2.2) | REF |
| Adjusted HR (95% CI) | 2.4 (1.5, 3.7) | 1.5 (0.98, 2.4) | REF |
| **J&J/Janssen** |  |  |  |
| n (%) | 317 (65.6) | 80 (16.6) | 86 (17.8) |
| Number (%) infected | 22 (6.9) | 3 (3.8) | 5 (5.8) |
| Crude HR (95% CI) | 2.1 (0.78, 5.5) | 0.98 (0.23, 4.1) | REF |
| Adjusted HR (95% CI) | 2.2 (0.82, 5.8) | 0.91 (0.22, 3.8) | REF |
| **Moderna** |  |  |  |
| n (%) | 124 (24.6) | 242 (48.0) | 138 (27.4) |
| Number (%) infected | 6 (4.8) | 6 (2.5) | 7 (5.1) |
| Crude HR (95% CI) | 2.9 (0.97, 8.7) | 1.0 (0.35, 3.1) | REF |
| Adjusted HR (95% CI) | 3.2 (1.0, 10.1) | 1.2 (0.38, 3.5) | REF |
| **Pfizer-BioNTech** |  |  |  |
| n (%) | 574 (45.5) | 387 (30.7) | 300 (23.8) |
| Number (%) infected | 24 (4.2) | 30 (3.8) | 33 (11.0) |
| Crude HR (95% CI) | 1.8 (1.0, 3.1) | 1.7 (1.0, 2.8) | REF |
| Adjusted HR (95% CI) | 2.0 (1.1, 3.6) | 1.8 (1.1, 2.9) | REF |

HR: Hazard Ratio, CI: Confidence Interval, REF = the >800 AU/mL group was considered the reference category in these models Models were adjusted for age (years), months since last vaccination dose, and SARS-CoV-2 infection before serologic test.

**Figure 2.**
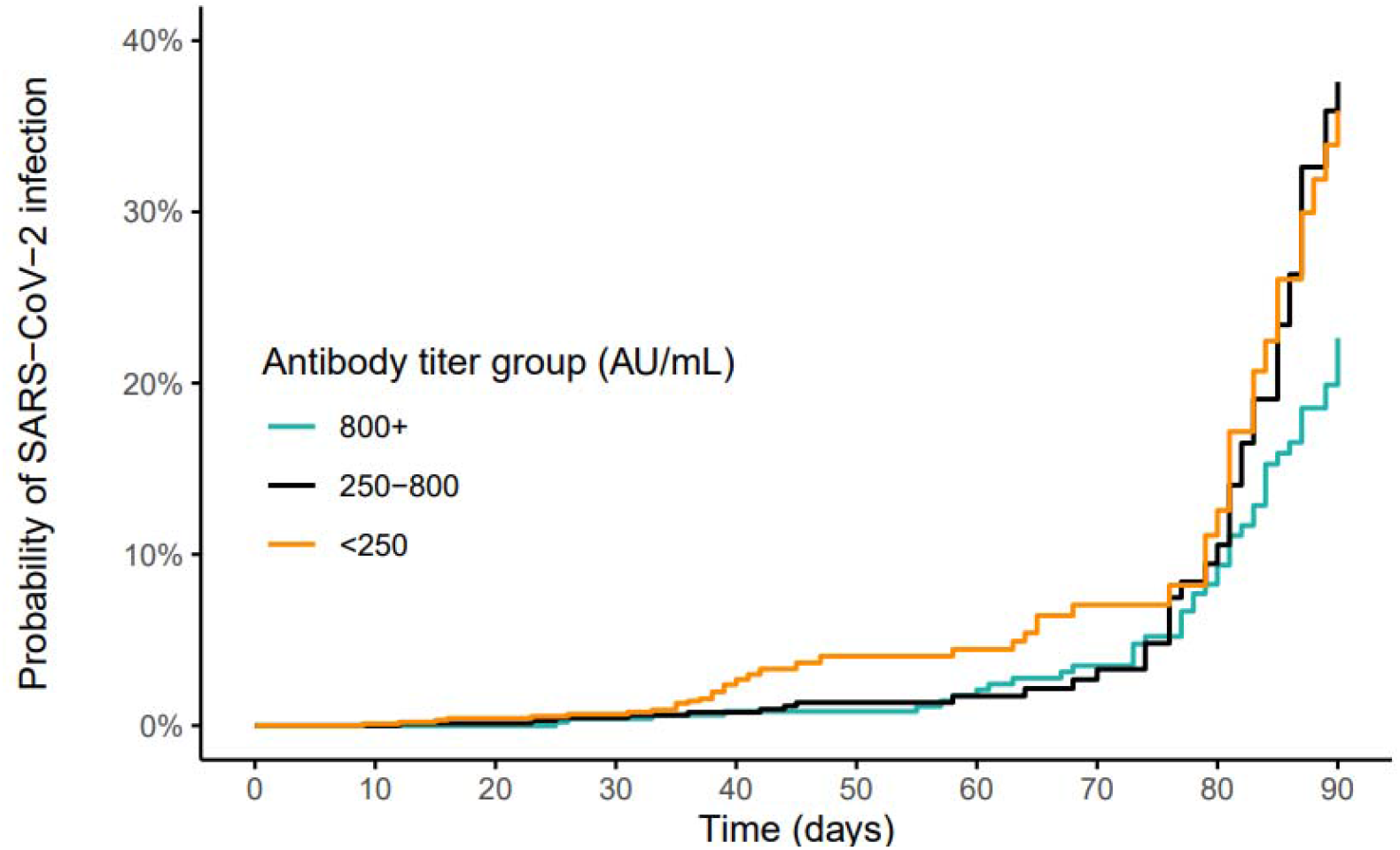
Time to infection by TrimericS IgG antibody titers. Figure 2 depicts the crude probability of SARS-CoV-2 infection within the 90-day follow-up period by titer group. Each step up indicates a new infection.

Among J&J/Janssen recipients individuals with titers <250 AU/mL had a higher relative rate of infection than those with >800 AU/mL (adj HR: 2.2; 95% CI: 0.82, 5.8), though not statistically significant.

Sensitivity analyses using other thresholds such as <125, 125-249, <200, and >750 AU/mL were also evaluated but did not meaningfully change results (Supplemental Table 1). Results were also robust to varying follow up periods as demonstrated by a 60-day analysis that showed a similar relative rate of infection comparing individuals with titers <250 and >800 AU/mL (adj HR: 2.8; 95% CI: 1.2, 7.0). In this shorter follow up period, the rate of infection was more similar between those with titers 250-800 and >800 AU/mL (adj HR: 0.94; 95% CI: 0.35, 2.5; Supplemental Table 2).

Individuals with titers <250 AU/mL were more likely to experience SARS-CoV-2 infections earlier in the follow-up period while Delta was predominant, comprising 67.3% of infections among this titer group (Supplemental Table 3). In contrast, 79.5% and 88.9% of infections among those in the 250-800 and >800 AU/mL groups, respectively, were from the Omicron variant. Notably, there were no Delta infections among individuals with titers >800 AU/mL; thus, the rate of Delta infections could not be estimated for this group. Individuals with titers <250 had a significantly higher relative rate of Delta infection compared to the 250-800 AU/mL group (adj HR: 7.0; 95% CI: 2.6, 18.9). Results from the variant-specific model for Omicron indicated that individuals with titers of 250-800 AU/mL had a significantly higher relative rate of infection compared to those with >800 AU/mL (adj HR: 1.8; 95% CI: 1.1, 2.9). The rate of Omicron infection among individuals with titers <250 AU/mL was not significantly different than those who were >800 AU/mL (adj HR: 1.2; 95% CI: 0.59, 2.3; Supplemental Table 2).

## Discussion

In this cohort of 2,248 vaccinated individuals, low antibody titers were associated with a significantly increased rate of SARS-CoV-2 infection in the subsequent 90 days. Across all titer groups, the rate of infection increased rapidly after day 70, which, for most of the cohort, coincided with the beginning of the Omicron wave for the US in December 2021. While the relative rate of Omicron infection among individuals with titers >800 AU/mL was lower than those with lower titers, they were more likely to experience an infection during the Omicron wave. This may be due to a decrease in antibody titers over the follow-up period, such that individuals with high titers at the time of serologic testing may have had low titers at the time of the Omicron wave, making them more susceptible to infection regardless of the variant. And those with low titers at the time of testing were more susceptible and likely to be infected earlier during the follow up period when Delta was predominant. Alternatively, as the Omicron variant is characterized by substantial immune escape compared to earlier variants [21,25–27], the antibody titers to the founder WA-1 strain as measured by the TrimericS assay may have had less relevance for Omicron than Delta. Previous reports have shown reduced correlation between measured anti-spike antibodies and neutralizing antibodies for Omicron infections when compared to the wildtype strain [15,27].

Vaccine-induced antibody levels wane over time, but earlier studies [28–30] assessed a relatively short window—generally up to 4 months post-vaccination. Since then, additional research [31–34] confirmed that antibody titers continue to decrease up to 6 months after SARS-CoV-2 vaccination and observed reduced neutralization activity to emerging variants including Delta and Omicron. Consistent with this prior research, the analyses in this study demonstrate that in a cohort of vaccinated individuals, titers decrease over time, as individuals who were vaccinated at least seven months prior to serologic testing tended to have substantially lower titers than those vaccinated within four months of testing.

This study has several limitations. Residual confounding by level of potential exposure may exist; for example, individuals with behaviors that result in higher risk of SARS-CoV-2 exposure may be more likely to have higher antibody titers and may also be more likely to be infected, this could lead to attenuation in observed associations. Our findings are based on a single antibody timepoint; multiple longitudinal datapoints could better characterize the trajectory of antibody decline following vaccination and quantify the level of immunity leading up to the time of infection. A large proportion of individuals (86.0%) were censored during follow up upon receipt of booster with 33% censored within the first 60 days. Sensitivity analyses showed that results from a 60-day follow-up period analysis did not differ greatly from the primary analysis with a 90-day follow-up period and demonstrated a slightly stronger association between titers and infection risk. Additionally, the proportion of censored individuals was similar across all three titer groups suggesting that this censoring did not introduce substantial selection bias. As this is a largely healthy, young, mostly male cohort, these results may not generalize to other populations, underscoring the need for further studies in more diverse cohorts. Finally, the results from these analyses may be specific to the timeframe observed during the end of the period of Delta variant dominance in late 2021 and early in the Omicron wave of 2021-2022 and thus may not generalize to other time periods during the pandemic or to other SARS-CoV-2 variants. Specifically, while high antibody titers are expected to predict lower risk of infection in most settings, the precise relationship between antibody titer and the risk of infection will depend on the antibody assay used, population, and epidemiological setting.

Answers to the question, ‘when should one get a SARS-CoV-2 booster dose?’ remain uncertain. Recommendations currently attempt to account for individual-level risks through proxy factors that include age, time since vaccination, vaccine type, and immunocompromised status [35]. Moreover, whether the logic for a Fall booster, like for seasonal influenza, will hold depends on (1) whether COVID-19 establishes a wintertime peak in transmission and (2) whether the duration of the immune response to the booster matches the duration of COVID-19 transmission. In contrast, findings from this study suggest that serologic testing can inform individual-level assessments of susceptibility to infection and therefore may serve as a useful guidepost for decision-making on when to get a booster.

More research and larger, more representative studies are needed to define correlates of protection. Additionally, it will be important to better understand the durability of immunity, for example how long someone may remain in a high or low titer group, and whether the protective effect from infection remains robust across SARS-CoV-2 variants. Further, it will be important to understand the degree to which new serological assays will be needed as circulating variants change; for example antibody levels measured by tests developed against the wildtype strain (WA-1) spike protein have been shown to have weaker correlation with Omicron-specific neutralizing antibody titers than wildtype titers [27,36]. The utility of these correlates may also encourage diagnostics manufacturers to develop, and the FDA to authorize antibody tests that are specific to neutralizing antibodies rather than overall IgG levels.

## Data Availability

Data produced in the present study are available upon reasonable request to the authors if approved by research committee governing these protected occupational health data records.

## Acknowledgements

We thank Joseph Fauver (University of Nebraska Medical Center) for contributing his expertise in genomic surveillance; Tempus Labs for their sequencing support; the NBA Players Association, the NBA Team Physicians Association, and medical and athletic training staff for the collection of these data; and the analytic and operational teams at the NBA (David Weiss, Miheer Mhatre, Patrick Clifton, Peter Meisel, Rachel Davis, Kelly Hogan, Caroline Coughlan, and Anton Arellano) and IQVIA (Kristina Zeidler, Gabriel Farkas, Kendall Knuth, Madeline Johnson, Erin Johnson, Riju Shrestha, Rahul Gondalia, Kelly Brotherhood, Sarah Connolly, Radhika (Melody) Samant, Julie Griffith, Jim Dunn, Marcin Mazurek, Tiffany Koch, and Michael Booth) for their tireless work on the NBA COVID-19 monitoring program.

